# Ileal derived organoids from Crohn’s disease patients show unique transcriptomic and secretomic signatures

**DOI:** 10.1101/2021.05.27.21257584

**Authors:** Barbara Joanna Niklinska-Schirtz, Suresh Venkateswaran, Murugadas Anbazhagan, Vasantha L Kolachala, Jarod Prince, Anne Dodd, Raghavan Chinnadurai, Gregory Gibson, Lee A. Denson, David J. Cutler, Anil G. Jegga, Jason D. Matthews, Subra Kugathasan

**Affiliations:** Division of Pediatric Gastroenterology, Department of Pediatrics, Emory University School of Medicine & Children’s Healthcare of Atlanta, Atlanta, GA, USA; Department of Biomedical Sciences, Mercer University School of Medicine, Savannah, GA, USA; Department of Biology, Georgia Institute of Technology, Atlanta, GA, USA; Division of Pediatric Gastroenterology, Hepatology, and Nutrition, Cincinnati Children’s Hospital Medical Center and the University of Cincinnati College of Medicine, Cincinnati, OH, USA; Department of Human Genetics, Emory University, Atlanta, GA, USA; Division of Biomedical Informatics, Cincinnati Children’s Hospital Medical Center, Department of Pediatrics, University of Cincinnati College of Medicine, Cincinnati, Ohio, USA

**Keywords:** Inflammatory bowel disease, Human Intestinal Organoids, Children, Epigenetics, Intestinal Epithelium, Secretome

## Abstract

**Background:** We used patient derived organoids (PDOs) to study the epithelial-specific transcriptional and secretome signatures of the ileum during CD with varying phenotypes to screen for disease profiles and potential druggable targets.

**Methods:** RNA sequencing was performed on isolated intestinal crypts and 3-week-old PDOs derived from ileal biopsies of CD patients (n= 8 B1, inflammatory; n= 8 B2, stricturing disease) and non-IBD controls (n= 13). Differentially expressed (DE) genes were identified by comparing CD vs control, B1 vs B2, and inflamed vs non-inflamed. DE genes were used for computational screening to find candidate small molecules that could potentially reverse B1and B2 gene signatures. The secretome of a second cohort (n= 6 non-IBD controls, n=7 CD; 5 non-inflamed, 2 inflamed) was tested by Luminex using cultured organoid conditioned media.

**Results:** We found a 90% similarity in both the identity and abundance of protein coding genes between PDOs and intestinal crypts (15,554 transcripts of 19,900 genes). DE analysis identified 814 genes among disease group (CD vs non-IBD control), 470 genes different between the CD phenotypes, and 5 FDR significant genes between inflamed and non-inflamed CD. The PDOs showed both similarity and diversity in the levels and types of soluble cytokines and growth factors they released. Perturbagen analysis revealed potential candidate compounds to reverse B2 disease phenotype to B1 in PDOs.

**Conclusion:** PDOs are similar at the transcriptome level with the in vivo epithelium and retain disease-specific gene expression for which we have identified secretome products, druggable targets and corresponding pharmacological agents. Targeting the epithelium could reverse a stricturing phenotype and improve outcomes.

## Introduction

Crohn’s disease (CD) is a main form of inflammatory bowel disease (IBD), a heritable chronic inflammatory disorder that can affect the entire gastrointestinal tract. Over 200 genetic susceptibility loci have been found to be associated with CD, of which 90% of the genetic associations are not in the protein coding regions, and heritability explained by only 15% of affected people having a first-degree relative.^1^ For many patients, suppression of inflammation by anti-TNF therapies is currently the primary treatment for CD. However, there is emerging data to support that anti-TNF therapies are effective in controlling the inflammation but not effective in preventing the development of a stricturing phenotype or disease progression, which are the two main complications of CD that often require surgical resection.^2^ Even newer biologic therapies offer relief in only a minority of patients. Thus, there is an urgent need for the development of newer treatments targeting different pathways and cell types.

The intestinal epithelium likely plays a pivotal role in the development and disease progression in CD, although the exact mechanisms are not clear.^3, 4^ By providing a physical barrier, protecting against foreign microbiota, aiding in nutrient and water absorption, hormone secretion, antigen uptake, and being a mediator of cyto/chemokine signaling,^5^ any defect in epithelial cell barrier function could have a disastrous consequence on homeostasis. The advent of intestinal organoids from individual patients (patient derived organoids - PDOs) allows for the growth of patients’ cells in culture from endoscopically derived mucosal biopsies^6^ and has revolutionized IBD research. Intestinal PDOs are three-dimensional structures grown in cell culture that are derived from multipotent epithelial stem cells residing at the base of intestinal crypts. They appear to retain many of the features and functions of the intestinal crypts/epithelium, such as supporting self-renewal, self-organization, barrier function, and differentiation into epithelial cell subtypes (Paneth, Goblet, transit amplifying, etc.).^7^ Hence, they provide major advantages compared to epithelial tumor cell lines, primary biopsies, or even animal models.

Herein, we sought to further define the similarities between organoids and intestinal crypts, and to test whether CD-specific transcription signatures are retained in PDOs after a short culturing time of 3 weeks, and if an epithelial specific secretome could be measured and used to draw conclusions about disease status. Recently published IBD data suggests that PDOs may lose their inflammatory signature after 5-6 passages,^8^ and thus we sought to investigate an earlier time point. To this end, we analyzed PDO transcriptomes from CD (both stricturing and inflammatory) as well as non-diseased healthy controls and used the changes in these transcription patterns to bioinformatically determine druggable targets for which known pharmacological compounds are available that could potentially reverse these CD phenotypes. Our results show that CD-specific transcript signatures are detectable in organoids and suggests that functional changes in the epithelium are potential drivers for CD onset and persistence as observed by changes in soluble cytokine and growth factors produced and released by patient derived intestinal organoids.

## Methods

### Patient selection and biopsy collection

Pediatric patients with Crohn’s disease were recruited for this study after obtaining consent during routine colonoscopy at the Children’s Healthcare of Atlanta / Emory University Hospitals from June 2019 through September 2019, and a second cohort collected between December 2019 and March 2021. Further, the patients undergoing endoscopy for abdominal pain but with grossly normal endoscopies and mucosal biopsies showing normal histology were included as non-IBD controls. In total, 29 mucosal biopsies from 16 CD patients and 13 non-IBD controls were obtained from the terminal ileum by colonoscopy. **Table 1** shows patient cohort characteristics used for RNA-seq analysis. **Supplemental Table 7** contains information on the cohort used in the secretome study. Study design and protocols were approved by the Emory University IRB committee. All authors had access to the study data and reviewed and approved the final manuscript. Patients with an established diagnosis of IBD undergoing routine colonoscopies, as well as newly diagnosed CD, were included in the study. Patients with UC (ulcerative colitis) or IBD-U (inflammatory bowel disease - undetermined) were excluded from the study. CD disease phenotype was determined by Montreal classification with divisions of B1 (n=8, non-stricturing, non-penetrating) and B2 (n=8 stricturing). Patients with B3 (penetrating) phenotype were not included in this study. CD samples were further classified into grossly and microscopically inflamed (n=**7)** versus non-inflamed (n=9).

**Table 1.**
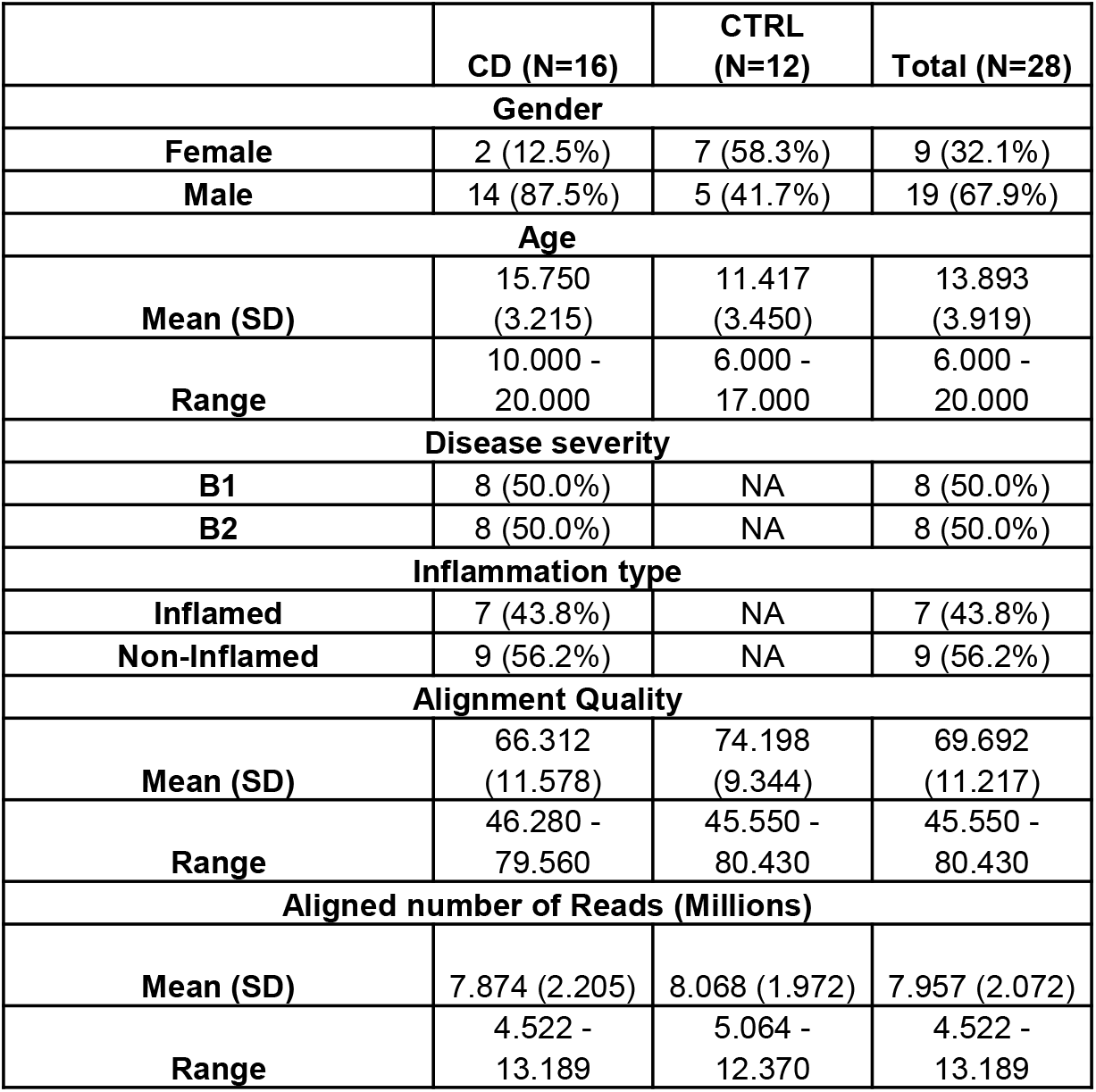

### Organoid culture

Two biopsy samples from the terminal ileum per individual were processed. Each sample was exposed to 15mM EDTA for 45 min at 4°C to aid with dissection. Biopsies were manually dissected under stereo microscopy to extract ileal crypts. Twenty percent of the crypt prep was used for RNA sequencing (stored in RNAlater at -80°C until RNA extraction), and the remaining 80% was used to initiate enteroid cultures. Samples were embedded in Matrigel and overlaid with culture media (50% Wnt, 20% R-spondin, 10% Noggin conditioned advanced DMEM/F12 media that included 100 ng/mL EGF, 10 µM SB202190, 10 nM Leu(15)-Gastrin-1, 2.5 µM CHIR99021, 0.5 µM A83-01, 2.5 mM Nicotinamide, 1 mM N-acetylcysteine, 10 µM Y-27632). Media was changed every 2-3 days for 3 weeks and passed at least twice before harvesting for RNA extraction. Media was collected on the third day if used in secretome studies and stored at -80C.

### Immunofluorescence

Organoids grown for 2 or more passages were removed from Matrigel with cold 4% formaldehyde/PBS and fixed for 15 min, washed and then permeabilized with 0.5% Triton X-100/PBS for 15 min. Organoids were maintained at room temp. in a liquid suspension within a microfuge tube during permeabilizing and staining, with gentle agitation every 5 to 10 min, and extensive washing between steps (buffer exchanges were performed by a brief centrifugation at 50 x g to settle organoids and the supernatant removed). Fixed and permeabilized organoids were blocked with 3% BSA/PBS and then incubated with primary antibodies (1/250 dilution in blocking buffer) for at least 2 hrs (E-cadherin, Sigma; Lysozyme, Dako), and next with secondary antibodies (1/1000 dilution in blocking buffer; goat, anti-rabbit 488 and -rat 647 Alexa Fluor conjugates, Thermo) and DAPI (1/10,000) (Thermo) for 1 hr. Stained organoids were mounted with Prolong (Thermo) between a microscope slide and coverslip and then imaged by confocal microscopy (Olympus FV100).

### RNA extraction and Sequencing

Total RNA was extracted from intestinal crypts and organoids using a Qiagen micro-RNA extraction kit and quantified by Nanodrop. RNA integrity numbers (RIN) were determined for samples on a bioanalyzer and only those samples with RIN values above 7 were sequenced. The library preparation was performed using QuantSeq FWD Kit; a protocol which is designed to generate Illumina compatible libraries of sequences close to the 3’ end of polyadenylated RNA. Libraries were sequenced using Illumina NextSeq 550 system by the Molecular Evolution core at Georgia Tech, USA.

### real time PCR

Total RNA from a subset of patients in **Table 1** (inflamed and non-inflamed CD) were tested. First strand cDNA was synthesized with 400 ng of RNA, oligo dTs and random primers in a final volume of 20 µL according to the manufacturer’s instruction (Thermo Scientific, USA). qPCR was performed as a 20 µL reaction with a final concentration of 1X TaqMan (Thermo Scientific, USA), 1 µL of Taqman primer, and cDNA. The reactions were carried out using single step real time PCR machine (Applied Biosystems) under the following conditions: initial denaturation at 95^0^ C for 5 min, followed by 40 cycles of 2 step amplification at 95^0^ C for 10 s and 60^0^ C for 30 s. Taqman primers SPINK4, HOXB2 and IGF2BP3, along with RSP01 as the internal control to normalize the RNA levels by delta-delta method, were all obtained from Thermofisher. Two technical repeats and experimental replicates were performed for each gene.

### Bioinformatics analysis

The sequenced raw FastQ files were aligned with reference genome hg38 using the STAR package^9^. In total, ∼11,500 protein-coding genes were considered with at least 10 read counts in 50% of samples. Differential expression (DE) analysis between disease status (CD cases against controls), among CD phenotypes (B1 against B2), and inflammatory status (CD inflamed against non-inflamed) groups were performed using Deseq2 package^10^ after adjusting for age and gender as covariates. The DE genes were identified with fold change (FC) ≥ 1.2 and false discovery rate correction (FDR<0.05) or nominal *P-*value < 0.05. *Pheatmap* was used to generate the heat maps for DE genes.^11^ The principle component analysis (PCA) was performed with the *prcomp* function and the PCA plots were generated using the *factoexra* package in R.^12^

**Pathway analysis** was performed with STRING^13^ and Panther^14^ pathway analysis using differential transcript lists and default settings.

### Perturbagen analysis

To discover potential small molecules likely to reverse B1 and B2 gene signatures, we used the LINCS cloud web tool from the NIH’s LINCS Library.^15^ We used the small molecule gene-expression signatures from the “Touchstone” dataset which has gene expression signatures on nine distinct cell lines after treatments, and includes over 8,000 perturbagens (>2000 small molecules including FDA-approved drugs).The LINCS cloud web tool compares queried signatures with gene expression profiles in the Touchstone library. Compounds whose signatures are anti-correlated to the queried signature are assumed to have a reversing effect, and hence may be of therapeutic potential, if the queried signature is from a disease state.

### Luminex

The used media from the organoid culture was analyzed for secretome diversity and levels. Supernatants of conditioned organoid media were collected 3 days after a fresh media change, followed by a brief centrifugation (500Xg for 5 min) and then stored at -80C. Subsequently, media supernatants were analyzed by magnetic bead-based multiplex LuminexTM assays for cytokines, chemokines, and growth factors, including FGF-basic, IFN-g, IL-12(p40/p70), IL-13, CCL5 (regulated on activation, normal T cell expressed and secreted or CCL5 [RANTES]), CCL3(MIP-1alpha), CXCL9 (monokine induced by gamma interferon or CXCL9 [MIG]), CCL4(MIP-1beta), VEGF, IL-1b, IL-2, IL-4, IL-5, IL-6, IL-2R, CCL2(MCP-1), CCL11(Eotaxin), IL-8, IL-10, IL-15, IL-17, IL-1RA, granulocyte-macrophage colony-stimulating factor [GM-CSF], granulocyte-colony stimulating factor (G-CSF), epidermal growth factor (EGF), HGF, TNF-a, IL-7, CXCL10(IP-10), and IFN-a (Human Cytokine 30-plex Panel, Life Technologies, USA), according to the manufacturer’s instructions using Luminex xMAP (multi-analyte profiling) technology. Results were plotted as picograms per milliliter.

## Results

### Similar expression profiles exist between freshly isolated intestinal crypt cells and 3-week-old PDOs

Manual dissection of ileal mucosal biopsies from pediatric patients yielded intestinal crypt preparations that were used for both RNA sequencing and generation of the organoid cultures (**Fig. 1A**), with typically several hundred organoids per biopsy being produced. Grossly, organoids derived from CD patients look like organoids derived from controls. To confirm the growth of epithelial-specific organoids in our small intestinal crypt culturing system, we immuno-stained for E-cadherin and lysozyme/Paneth cells (**Fig. 1B)**. In addition to staining positive for E-cadherin and lysozyme, these cultures also produced Goblet cells (MUC2 staining, data not shown). During the culturing of organoids, we observed few, if any, differentiating morphological characteristics between CD and controls after 3 weeks of culture with at least 2 passages (**Fig. 1C**). Occasionally, severely inflamed, and friable tissue from CD patients would produce fewer crypts yielding poor growth and subsequent culture failure (∼10% of CD cases).

**Fig. 1.**
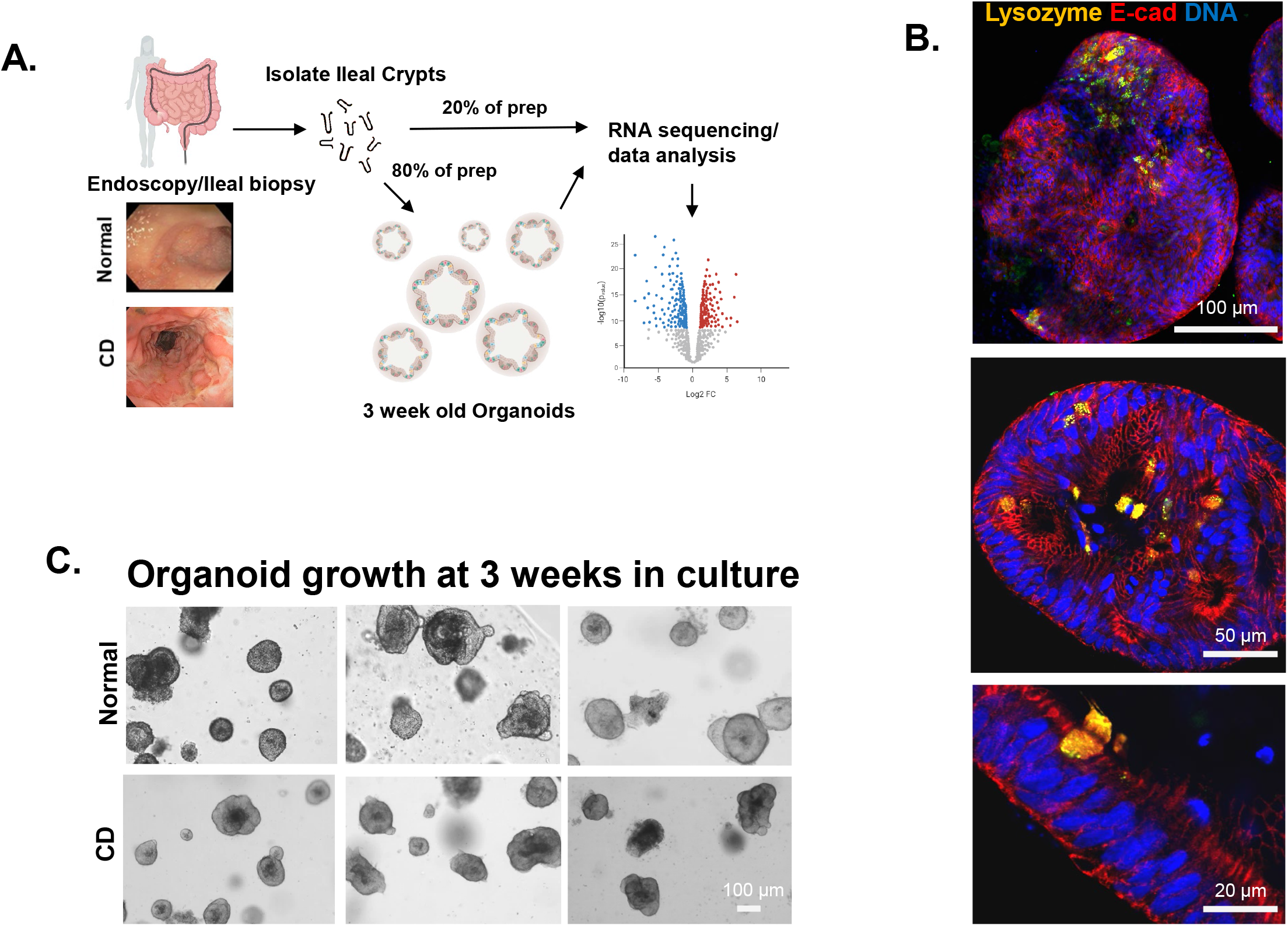
Consort diagram of the study and processing of small intestinal crypts/organoids. (A) Schematic representation of the study shows the workflow of biopsy to crypt isolation, organoid culture, RNA extraction and transcriptomic data analysis. (B) Immunofluorescence staining of small intestinal organoids for lysozyme (gold), E-cadherin (red), and DAPI (blue) showing epithelial-specific cell types. (C) Microscopic images of mature organoids in culture at 3 weeks are shown for both CD patients and non-IBD controls.

We compared the entire transcriptomic profile (19,900 protein coding genes from hg38), between ileal crypts and corresponding organoids. Of those, we detected 15,554 genes expressed in either ileal crypts and/or organoids, and 90.9% of those genes (n=14,149) were expressed in both organoids and crypts. Around 7.0% (n=1,091) of the genes were expressed only in ileal crypts, and the remaining 2.0% (n=314) genes were expressed only in PDOs. We noticed a strong positive correlation in mean expression levels with R^2^ = 0.906; *P* < 10^−16^ (**Fig. 2A**) for the genes that are expressed in both ileal crypt and PDOs. As depicted in **Fig. 2A-B**, the remarkably similar expression between PDOs and ileal crypt provides evidence that the PDOs are a suitable epithelial model to test druggable targets or measure IBD-specific molecular signatures. Pairwise comparative analysis on entire transcriptomic data of CD and non-IBD controls obtained from both ileal crypts and organoids showed similar patterns between the samples (**Supp. Fig. 1**), consistent results with **Fig 2A**. Next, we further explored 1,091 genes that were expressed only in the ileal crypts using STRING^13^ pathway analysis. This revealed a set of genes that encode proteins involved in immune system signaling/regulation and/or GPCR signaling (**Supp. Fig. 2**). The expression of several protein coding genes involving cellular adhesion were also detected only in the intestinal crypt. Similar analysis for the same genes in Panther pathway^14^ revealed the three most prominent pathways represented from the “crypt-only” data were 1) inflammation mediated by chemokine and cytokine signaling, 2) heterotrimeric G-protein signaling pathway-Gq alpha and Go alpha pathway and 3) Wnt signaling.

**Fig. 2.**
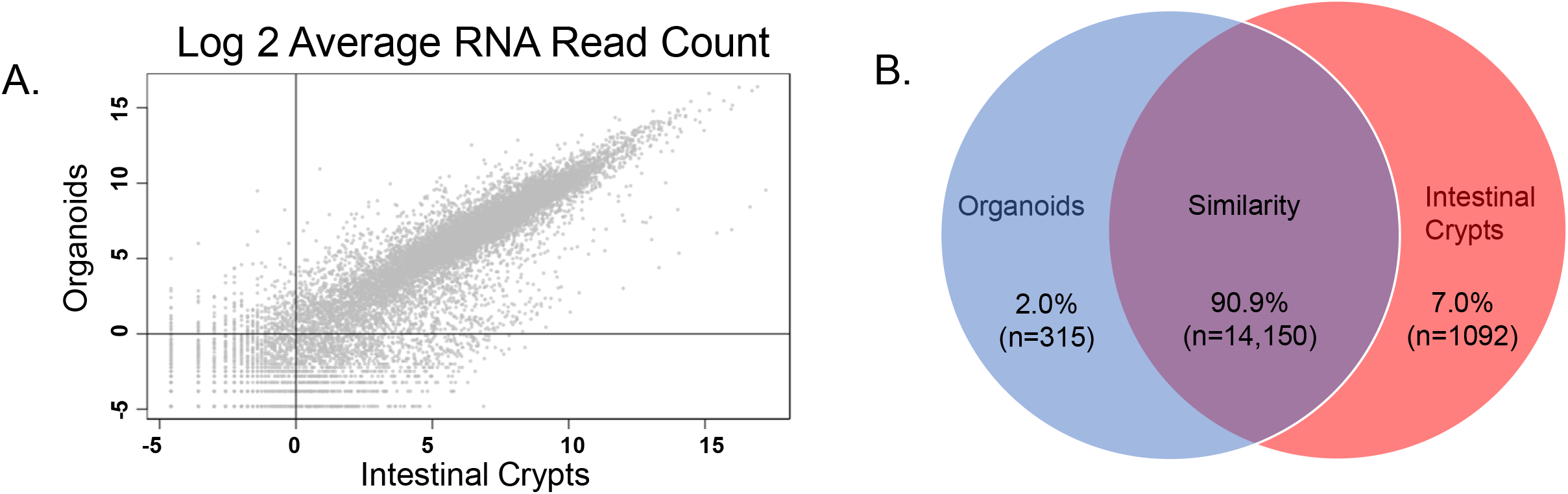
Transcriptomic signatures reveal epithelial nature in patient derived organoids. (A) Scatter plot shows the protein coding genes are compared between organoids (X-axis) and ileal crypts (Y-axis). Both X- and Y- axis shows the log transformed average read counts. (B) Venn diagram shows the number of genes expressed in both organoids and ileal crypts (purple), genes that are expressed only in ileal crypts (blue), and genes that are expressed only in PDOs (red).

### Disease specific transcriptomic changes within PDOs

To examine more subtle differences than gross morphology, we performed differential expression (DE) analysis on bulk RNA sequence data from samples processed as described in **Fig. 1**, comparing CD organoids to non-IBD control organoids. The analytical representations of the interrogated data sets in PC plots, volcano plots and heat maps of significantly different transcripts are shown in **Fig. 3A-C, Supp. Table 1**. The first two principal components (PCs) from the entire transcriptomic profile in PDOs show separation of CD and non-IBD controls along the axis of PC2 (*P* = 0.0046) (**Fig. 3A**). The clustering indicates that the PDOs retained a CD-specific gene signature. We performed DE analysis among the disease groups (CD vs non-IBD controls) in PDOs and identified 817 genes to be significantly different at *P* < 0.05 (**Fig. 3B**). Of the 819 genes, 393 were increased in CD (up-regulated) while 426 were decreased (down-regulated). Consistent with the PC analysis in **Fig. 3A**, except for a few CD cases, the hierarchal clustering heatmap identified two independent clusters for CD and non-IBD controls using DE genes (**Fig. 3C**). Transcripts encoding several soluble factors were also of interest (**Supp. Fig 3**). Further, Panther pathway analysis revealed 298 pathway hits, with the top three pathways being 1) Wnt signaling, 2) inflammation mediated by chemokine and cytokine signaling, 3) Gonadotropin-releasing hormone receptor pathway. Collectively, our results indicate that the CD-specific transcriptomic profiles were retained in the PDOs after several weeks in culture. It appears that these gene signatures correlate with pathways occurring in non-epithelial cells whose growth is not supported by our culturing conditions. This suggests that disturbances in the crosstalk between non-epithelial mucosal cell types and the epithelium are taking place during IBD, and thus PDOs are reflecting retained defects of the epithelial response to altered extracellular stimulation taking place in the mucosa during CD.

**Fig. 3.**
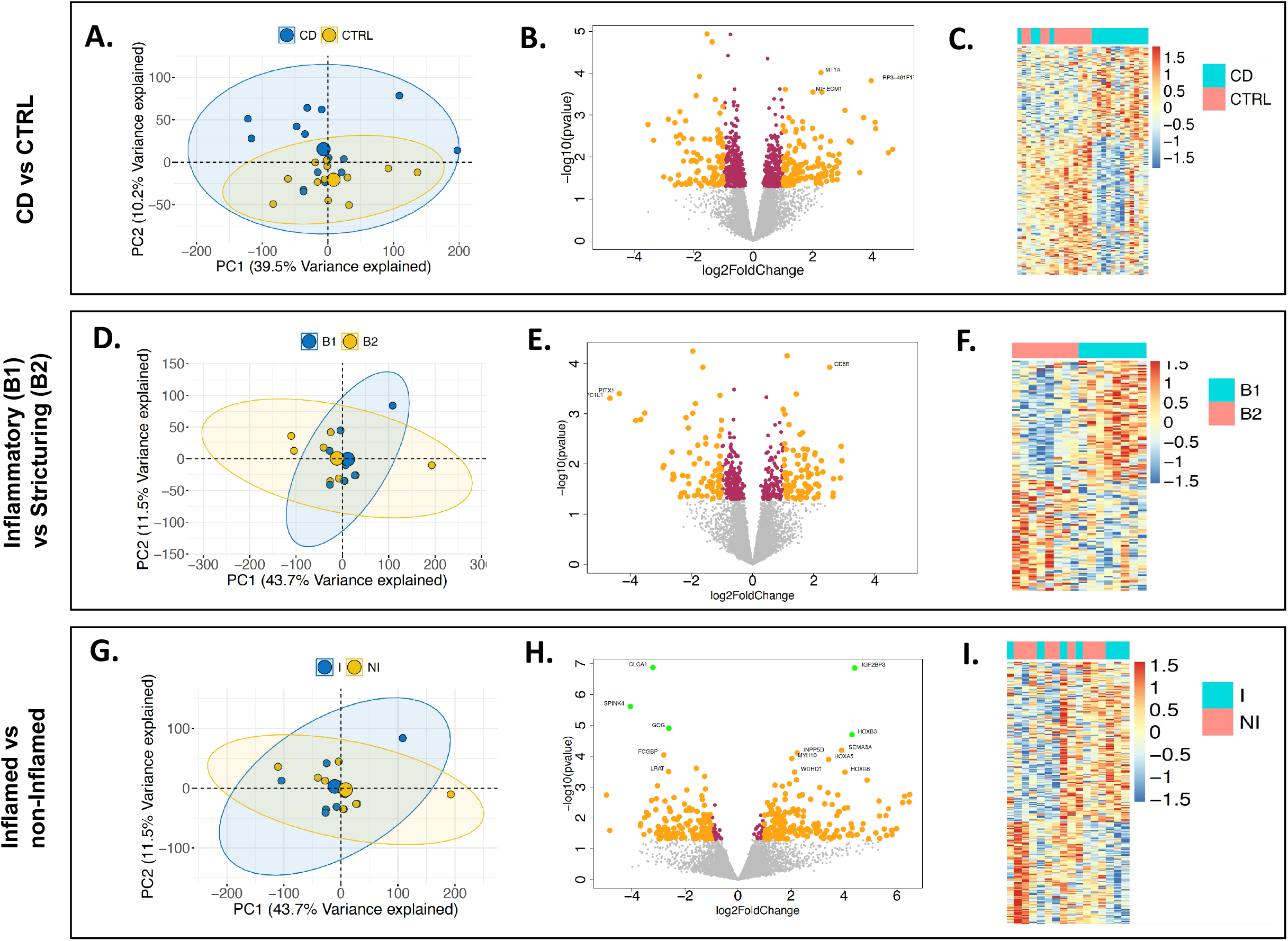
Transcriptomic signatures in PDOs retain disease states. (A) Principal component analysis (PCA) plots with entire transcriptomic data revealed disease specific expressions (CD vs non-IBD controls). (B) Volcano plot representation on differential expression (DE) results between CD and non-IBD controls show both up- and down- regulated genes. (C) Hierarchical clustering on DE genes shows two independent clusters for CD and non-IBD controls. (D) PCA plots with entire transcriptomic data within CD patients not showing any clusters among disease phenotype (B1 and B2). (E) DE analysis between B1 and B2 showed transcriptomic signatures specific to disease phenotypes. (F) Hierarchical clustering on disease phenotype DE genes showed two independent clusters. (G) PCA plots with entire transcriptomic data among Inflammation status (Inflamed and non-Inflamed), within CD patients. (H) DE analysis between Inflamed and non-Inflamed showed transcriptomic signatures specific to Inflammation status. (I) Hierarchical clustering on Inflammation status specific DE genes showed two independent clusters. In PCA plots, the first two principal components are plotted along with their variance explained. In Volcano plots, the log2 fold changes, log transformed *P* values are shown on the X- and Y- axis, respectively. Maroon shows the DE genes with P < 0.05 and yellow shows P < 0.05 and FC >1.2.

### Phenotype specific changes in PDOs from CD

We recently showed the importance of cellular crosstalk in IBD by predicting the progression to stricturing (B2) or penetrating disease (B3) within 3 years of diagnosis ^16^. The addition of transcriptomic signatures from bulk RNAseq data generated from whole ileal biopsies into the analysis significantly improved the accuracy of the risk score. Furthermore, many of the transcription signatures observed to be enriched during B2 in that study belonged to genes involved in extracellular matrix production, most of which are generated by subepithelial myofibroblasts and another non-epithelial cells resident within the lamina propria. Given our pathway analysis suggesting potential changes in gene expression for soluble molecules such as cytokines, chemokines and growth factors that regulate the mucosal cellular interactions between the epithelium and lamina propria cells, we further investigated the transcriptional differences that might be contributing to an increase in fibrosis. The use of intestinal PDOs in a growth window where they are still retaining their CD-specific transcription signature (3 weeks) allowed for strict observation of the transcriptional changes within the epithelium occurring during disease progression early on from B1 to B2 status (**Fig. 3D-F, Supp. Table 2**). The PC analysis of the entire transcriptomic profile between B1 and B2 patients did not significantly separate B1 from B2 disease phenotypes (**Fig. 3D**), and the DE analysis between B1 and B2 showed CD phenotype specific differences in only 470 of 11,933 protein coding genes with p <0.05 significance, namely fewer than expected by chance. Nevertheless, 156 of these genes had a FC > 2 (absolute difference in log2 expression >1), with 83 up-regulated and 73 down-regulated in B1 cultures (**Fig. 3E)**. This results in two distinct clusters in the heatmap hierarchal clustering for B1 and B2 CD phenotypes using DE genes (**Fig. 3F**). Panther pathway analysis on these DE genes identified 188 hits and the top three pathways between B1 and B2 were the same between CD vs control PDOs but differing in ranking order 1) gonadotropin-releasing hormone receptor pathway, 2) inflammation mediated by chemokine and cytokine signaling, 3) Wnt signaling. Thus, the change in chemokine/cytokine differences between B1 and B2, though not formally significant given the sample size, nevertheless might be related to the onset of signaling leading to changes in extracellular matrix production and the fibrosis observed later in B2/B3 transition.

### Inflammation specific changes in PDOs

Finally, we looked at whether the PDOs transcriptomic profiles were different within CD patients with respect to inflammatory status (inflamed vs non-inflamed) in the mucosa at endoscopy (**Fig. 3G-I, Supp. Table 3**). The PCs from the entire transcriptomic profiles do not classify the inflammatory status in the PDOs (**Fig. 3G**). Despite just 358 of 11,933 genes being differentially expressed at p < 0.05, the fold changes for this comparison in **Fig. 3H** are much greater than in the above comparisons, suggesting considerable heterogeneity in the expression profiles of the inflamed crypt preparations. A set of five genes at or close to experiment-wise significance (p<4×10^−6^) were significant by FDR < 0.05, including increased expression of *IGF2BP3* and *HOXB3* in inflamed PDOs, while *CLCA1, SPINK4* and *GCG* were decreased. The hierarchal clustering of DE genes in the heatmap confirmed the substantial heterogeneity between the two groups (**Fig. 3I**). Panther pathway analysis on DE genes identified 121 hits, with the PDGF signaling pathway being the most represented within the data, followed by two pathways with equal ranking for second highest (angiogenesis and CCKR signaling), followed by seven pathways in third of which dopamine receptor mediated signaling, inflammation mediated by chemokine and cytokine signaling, and integrin signaling were included. We used a second method to confirm these RNA-seq observations, and the qPCR assays using Taqman primers for SPINK4, IGF2BP3, and HOX2B (**Suppl Fig 4**) substantiated these findings.

**Fig. 4.**
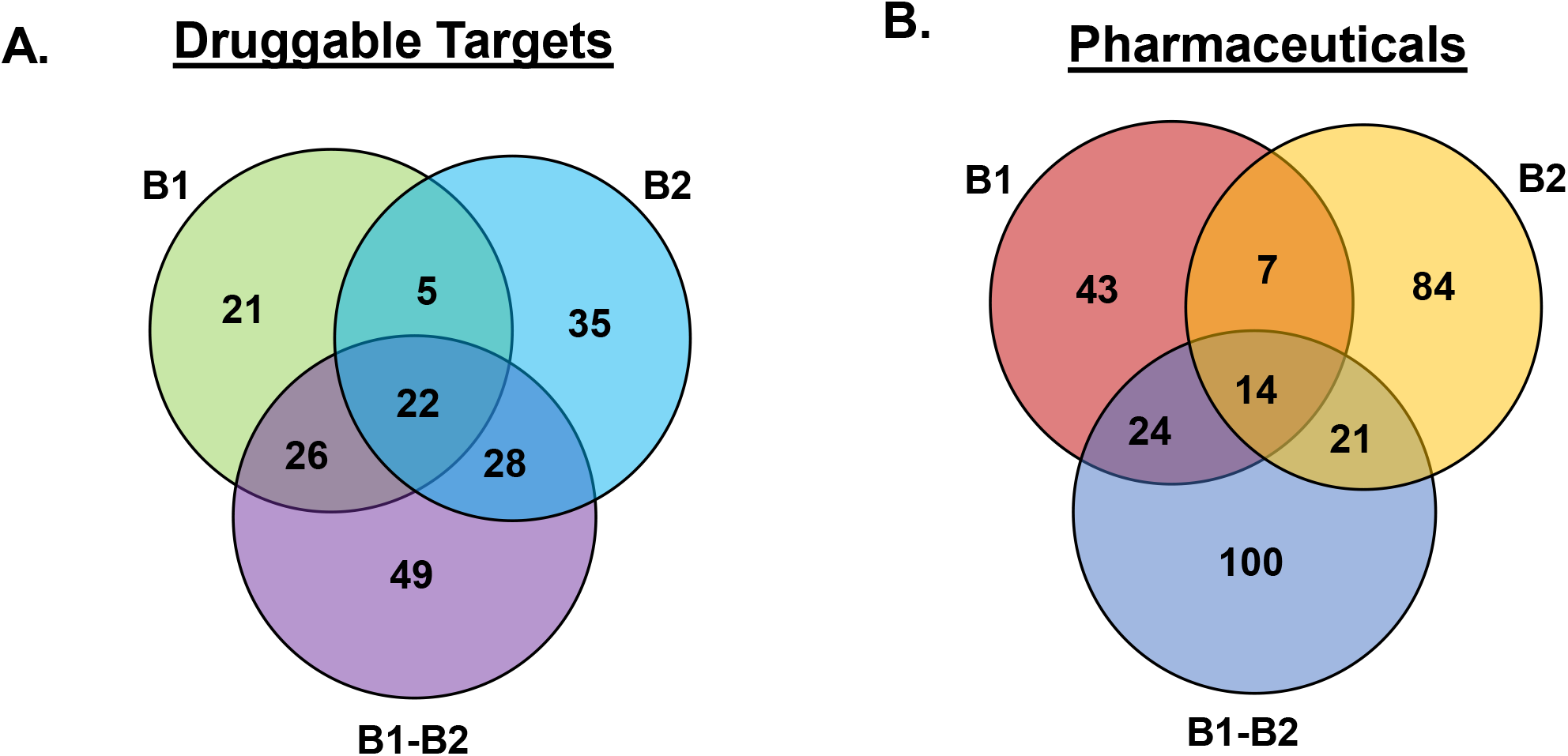
Drug classes and small molecules for B1 and B2 DE genes. (A) The drug classes (mechanism of action) identified for B1 (B1 vs CTRLs), B2 (B2 vs CTRLs), and shared genes among B1 and B2 DE genes. (B) Perturbagen analysis identified small molecules (approved or investigational compounds) belonging to the drug classes outlined in A.

### Small molecules likely to reverse B1 and B2 gene signatures

We asked whether it was possible to computationally predict, based on known small molecules (approved drugs and investigational compounds) transcriptomics data,^15^ candidate therapeutics that might potentially reverse epithelial specific gene signature observed in B1 and B2 organoids. To perform this, we used three sets of differentially expressed genes (DEGs) from B1 vs non-IBD controls (n=184 DEGs, **Supp. Table 4**), B2 vs non-IBD controls (n=323 DEGs, **Supp. Table 5**) and the common genes (n=57 DEGs, **Supp. Fig. 5**) that were consistently up-or down-regulated in both B1 and B2, when compared to controls. We tested all these genes independently against the Touchstone dataset^15^. The perturbagen analysis on the three DE gene sets identified 14 compounds common to all (**Supp. Fig. 6**). The compounds identified for each DE list are provided in **Supp. Table 6**. Pathways targeted by the 14 compounds include: Rho kinase inhibitor, PI3K inhibitor, interleukin receptor agonist, cholecystokinin receptor antagonist, etc. (**Supp. Fig. 7)**. Overall, our analysis identified 14 potential small molecules for 25/32 genes that represent the core signature (of B1 and B2), reversal of which could be broadly therapeutic. The summary score reflects the ability of a given small molecule to reverse a disease-specific gene signature. The number of candidate therapeutics mechanism of action (drug classes or categories) is shown in **Fig. 4A**, while the number of small molecules with the highest scores for therapeutic potential are shown in **Fig. 4B** and corresponding lists are shown in **Supp. Table 6**. Experimental validation as well as clinical evaluation will be required to establish whether computational prediction of drug targets using organoid gene expression profiles has presented a new avenue to potentially test the efficacy of old and new pharmaceuticals in reversing the damage of IBD.

**Figure 5.**
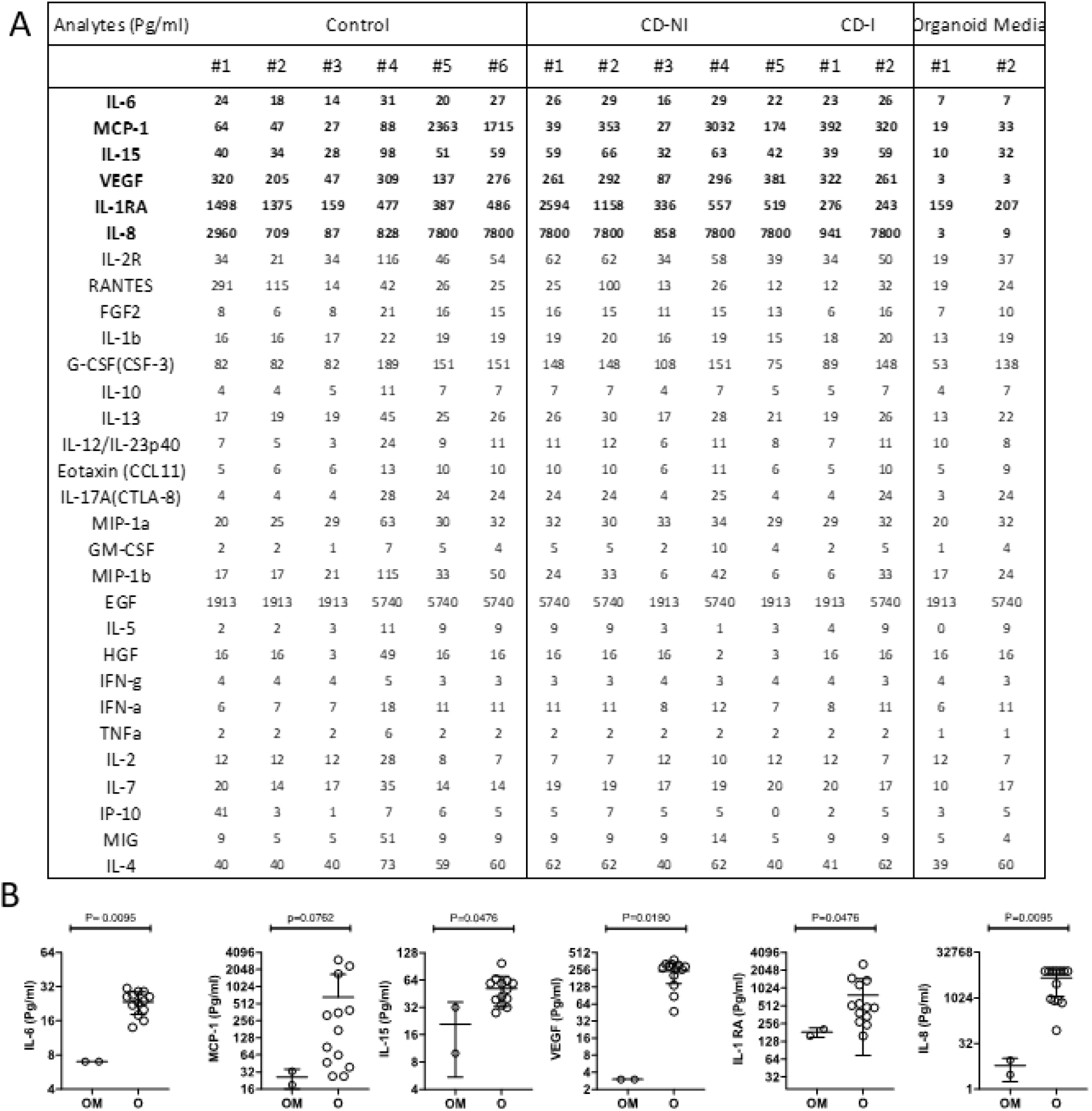
Secretome analysis of patient derived organoid conditioned media. Thirty-plex secretome analysis was performed on 3-day old organoid conditioned media. (A) Concentrations of 30 analytes from the supernatants of Control, Crohn’s (No inflammation; CD-NI) and Crohn’s Inflammatory (CD-I) organoids are shown. Analyte concentrations in fresh organoid media (background reference) is also shown. Highlighted analytes are statistically significant over the organoid media (background reference). (B) Six out of 30 analytes that are significantly higher in organoid condition media (O) over the background Organoid Medium (OM) is shown. Shown results are the cumulative of two independent Luminex 30-plex runs.

### Secretome profiles of ileal organoids

Given that the pathway analysis of transcriptional differences in the organoids pointed strongly towards changes in cytokines, growth factors and other soluble molecules released by epithelial cells, we next sought to determine the divesity and levels of secretome released from organoid cultures to further assess any disease associated characteristics of these samples at the protein level that might functionally impact the behavior of other cell types in the mucosa in a paracrine fashion during CD. The conditioned media from each organoid culture was collected from a second cohort of CD patients (n=6) and non-IBD controls (n=7, 2 inflamed, 5 non-inflamed, **Supp. Table 7**) and tested by Luminex with a 30-analyte panel (**Fig. 5**). The conditioned media from the organoid cultures contained very low levels for many of the analytes including TNF-alpha and IFN-gamma, even for the inflamed samples. The high levels of EGF were the result of its presence in the organoid media for growth purposes. Other molecules in the media produced by patient derived organoids that were readily detectable above baseline levels (we used fresh organoid media as a negative control for baseline determination) were IL-8 (at high levels), IL-15 (moderate levels), and FGF2 and IL-6 (at low levels), but not at statistically significant differences between the CD and controls. However, several of the analytes were detectable at highly variable levels across the samples, and the unique profiles of each patient sample across the 30 analytes appears to have a diagnostic potential. In **Figure 5**, MCP-1, VEGF, IL-8 and IL-1RA levels vary considerably across the patient samples and may indicate differences in epithelial health/function between the patients when taken into context with the production and levels of other secretomic signals.

## Discussion

Since the original work of Sato et al.^6^ organoid systems have expanded over the last decade allowing for genetic and functional ex-vivo studies by recapitulating many epithelial cell types and functionalities of the original tissue from which they are derived. The intestinal organoid model has propelled work in cancers such as colorectal, gastroesophageal, and prostate cancer, as well as gastrointestinal diseases such as inflammatory bowel disease and celiac disease.^17-19^ Herein, we evaluated the capability of pediatric CD PDOs to retain in-vivo and disease specific transcriptional patterns, evaluated how these features differ across phenotypes, and computationally predicted pharmaceuticals to target the CD-specific differences. We found remarkable similarity between in-vivo epithelium and organoids at the transcript level and observed that although CD organoids are not morphologically distinct from non-IBD controls, they do retain many disease-specific transcriptional changes that revealed epithelial-specific pathways, and show a diverse, patient-specific secretomic profile, any of which could potentially serve as diagnostic signatures or pharmaceutical targets.

While many studies have relied on organoids, few studies have compared the in vitro transcript signatures to that of the original in vivo epithelium. Middendorp et al. evaluated location specific intestinal transcriptional patterns in mice organoids and found crypt-derived genes being highly maintained in organoid cultures without location-specific external signals from mesenchyme or luminal content.^20^ Fuji et al. recently showed the similarities between fresh human intestinal crypts and organoids,^7^ however the profiles generated in that study were limited by analyzing only a few patient samples. We have expanded on the Fuji et al. study by including more patients, along with disease subtypes, and found consistent results. Our studies demonstrate that most genes expressed in the in vivo epithelium are also represented in the organoid cultures (**Fig. 2B**) and that the 7% of genes found in the intestinal crypt prep, but not organoids, are likely due to the retention of some immune and mesenchymal components during crypt extraction from mucosal biopsies. VanDussen et al showed that current culturing conditions for intestinal organoids derived from isolated crypt preps grow exclusively epithelial cells despite the initial culture having numerous cell types.^21^ However, as we have seen, some of these epithelial-specific gene signatures may be stimulated by mesenchymal/immune crosstalk in the mucosa and are lost during organoid culturing or are part of villi that are not reproduced in organoid cultures. Focusing on the genes found only in the intestinal crypt prep, the inability to detect *FPR1, CLDN5* and *8*, or *MUC4* in organoids, which are normally expressed in intestinal epithelial cells, suggests that some downregulation of genes occur in culture, a similar phenomenon also observed by Fuji et al.^7^ However, detection of expressed genes such as *CD4, CCL22, LY9, CCL18, TLR7* and *CCL3* suggest the presence of immune cells, while *RSPO3, MMP10* and *ADAM19* are indicative of stromal cells that were in the intestinal crypt prep after extraction from a mucosal biopsy. It is unclear how the expression of certain epithelial genes is down regulated in organoids upon culturing, but it is likely that the loss of crosstalk with other mucosal cell types plays a role. Indeed, Fuji et al. found that the addition of insulin like growth factor 1 (IGF-1) and fibroblast growth factor 2 (FGF-2) to the culturing media was required in order to create a transcription profile in the organoids that aligned even closer with that of fresh crypts.^7^ Thus, the lack of ligands in the culturing media that are normally produced by non-epithelial cells in the mucosa may drive the loss of cognate GPCR signaling pathways we observed in organoids during culturing (**Supp. Fig. 2**).

Although there were no clear morphological characteristics observed between CD and control organoids by gross microscopy (**Fig. 1C**), there were measurable differences at the level of transcript abundance (**Fig. 3A-C**). However, a recent report by Kaline et al. showed that after 1 week in culture PDOs of inflamed origin lost the majority of their inflammatory gene expression but after four weeks in culture (5-6 passages) the UC diseased group remained transcriptionally distinct from non-IBD controls.^8^ In contrast to those findings, UC organoids grown for only 1 passage (about 2 weeks) after extraction from mucosal biopsies showed significant differences at the transcript level when compared to controls.^18^ In that study, Dotti et al. investigated expression differences that persisted in epithelial organoid cultures generated from patients with UC as compared to non-IBD subjects and found a group of differentially regulated genes in UC patients associated with antimicrobial defenses, as well as secretory and absorptive functions.^18^ Taken together, it appears that many IBD-specific transcriptional changes are retained within organoids, at least for some period of time in culture, and these changes likely reflect defective signaling in the gastrointestinal epithelium, perhaps early and distinct, that plays an important role in both forms of IBD onset and progress. In our study, we found a strong signal for inflammatory chemokine and cytokine pathways associated with ileal organoids from CD patients. Finding that a repertoire of soluble factors is being released by the epithelium, some of which differ drastically between patients, substantiates this possibility. Although we did not find group-wide statistical differences between CD and control, the unique signatures for each patients sample found across the analytes, regardless of phenotype, points towards the disruption of crosstalk that can take place when unique variables are disrupted out of homeostatic alignment. Our method of epithelial-specific profiling of secretomes will allow for a better understanding of mucosal cellular crosstalk as we continue to develop this new protein pipeline and further refine our observations. Our current methodology shows promising potential in elucidating the relationship between gene expression, protein signaling, and cytokine release (secretome) specific to the intestinal epithelium.

In fact, of the 200 epithelial specific genes involved in the innate immune response, we found 5 of the gene transcripts to be markedly different between organoids from CD versus non-IBD patients (**Supp. Fig. 5**). *CXCL5* and *CCL5* transcripts were more abundant in CD, while *IL33, PDGFB and ILDR2* were found to be less abundant. *CXCL5* is a lipopolysaccharide induced chemokine, and an important attractant for immune cell accumulation. It has been studied in various cancers, and is increased during IBD, specifically UC patients, indicating a potential role of the epithelium in the regulation of leukocyte migration, general systemic inflammation and advancement of disease severity in IBD patients.^22, 23^ Zhang et al. recently reported *CXCL5* overexpression as predicting a poor prognosis in pancreatic ductal adenocarcinoma, while Nouailles et al. described pulmonary epithelial cells that were secreting *CXCL5* and driving progression of tuberculosis.^24, 25^ The increase in *CXCL5* and *CCL5* chemokine transcripts suggests that underlying inflammatory signaling initiated by the epithelium could be playing an important role in driving IBD pathogenesis^26^, and although we did not find direct correlation between mRNA levels and extracellular cytokine protein levels, both of these soluble proteins (CXCL5 and CCL5) were detectable in the media of patient derived organoids, along with a number of other pro- and anti-inflammatory cytokines, including IL-8 and IL-1RA, respectively, and the angiogenic factor VEGF.

Indeed, a predominate signature setting the groups of organoids apart was the differential transcription patterns that pointed strongly to immune and angiogenic pathways originating in the epithelium. For example, increases in *CCL5* transcript were up in CD vs controls and *CX3CL1* transcript was up in B1 vs B2. *CCL5*, also known as RANTES, is most commonly reported in HIV literature and cancers, and found to be increased in in both UC and CD.^26, 27^ Berrebi et al. also noted that in pediatric patients with Crohn’s disease, there was an increase of *CCL5* transcript in mucosa within epithelial and immune rich regions^28^, further indicating a role for this cytokine during IBD, and the epithelium as a likely source for its production. Remarkably, PDGF (platelet-derived growth factor) signaling differed the most between inflamed and non-inflamed samples, which was previously found to be a promising target for anti-fibrotic and anti-angiogenic therapies.^29^ The role of PDGF signaling in IBD is unknown, however it was increased exclusively in active IBD patients in a small study from Poland.^30^ We found our CD organoids as a whole exhibit a decrease in *PDGFB* transcript levels, and for inflamed samples, a decrease in *PDGFA* and *PDGFC* transcript levels, suggesting that the epithelium might also play a role in shaping angiogenic properties associated with IBD.^31^ The Luminex data supports this (**Fig 5**), as VEGF was released at differing levels, suggesting that angiogenic pathways are at play within the epithelum and may reflect changes in crosstalk during CD. While no differences were found in cytokine transcripts between non-inflamed vs inflamed, *GNG4* was in lower abundance in the inflamed group. Interestingly, *GNG4* is under heavy epigenetic control and is extensively hypermethylated during glioblastoma formation where its expression is downregulated leading to a negative impact on the function of GPCRs and chemokine receptors.^32^ Whether changes in *GNG4* methylation occur in the ileal epithelium during IBD has not been determined, but changes in DNA methylation do occur systemically with increases in inflammation.^33^ The Wnt pathway is also mediated by GPCR signaling,^34, 35^and changes in transcript levels encoding proteins involved in Wnt pathways were associated with B1 and B2 phenotypes further indicating defects in pathways requiring crosstalk with non-epithelial cells and the ligands that drive this signaling. Looking at ways to correct these phenotypes by targeting the most common intersection in physiological changes, our pathway and druggable target analysis found two molecules in particular, actarit and devazepide, an interleukin and CCK receptor blocker (CCK is a GPCR), respectively, that appear to have the most potential in correcting some of the defective signaling observed in the epithelium during CD (**Fig. 4/Supp. Figs. 5, 6, 7**).

Overall, our data supports PDOs sharing many features at the transcriptome level with the in vivo epithelium and retain disease specific phenotypic defects in gene expression. Our process shows that from two biopsies, we can generate several hundred PDOs in 2-3 weeks while retaining the disease specific phenotypes. These PDOs and the conditioned media they generate in culture can be used for further investigations into mechanistic studies of disease progression particularly structuring and fibrogenesis and potential drug screening. We have identified druggable targets and corresponding pharmacological agents, along with potential endocrine factors originating in the epithelium that may contribute to disease. Many of the pathways shown to be altered in CD are related to immune/non-epithelial crosstalk that is lost upon culturing organoids, suggesting that a change in the ability of the intestinal epithelium to perceive and transmit signaling with non-epithelial cells is taking place during CD. It is not clear on the cause of the release of IL-8, IL-1RA and VEGF or other cytokines/growth factors from the PDOs during culturing. Or whether these differences are measurably different to other patient samples that can better withstand deficiencies in extracellular cues lost during culturing organoids, and thus more experiments are required to unravel the function of the secretome in these processes. Therefore, future work will begin to interrogate the functional consequences of these differences in transcriptome and secretome, and the effects they have on the epithelium and non-epithelial cells in the mucosa.

## Data Availability

The data is available upon request to the corresponding author

## Abbreviations

(CD): Crohn’s disease
(DE): Differentially expressed
(DEGs): Differentially expressed genes
(FDR): False discovery rate correction
(FC): Fold change
(PDO): Patient derived organoids
(PDGF): Platelet-derived growth factor
(RIN): RNA integrity numbers
(PCA): Principle component analysis
(SRA): Sequence Read Archive
(UC): Ulcerative colitis

## Acknowledgements

We like to thank Shweta Biliya at GA Tech for preparing libraries and performing sequencing.

---

**Author names in bold designate shared co-first authorship**.

**Supplemental 1.**
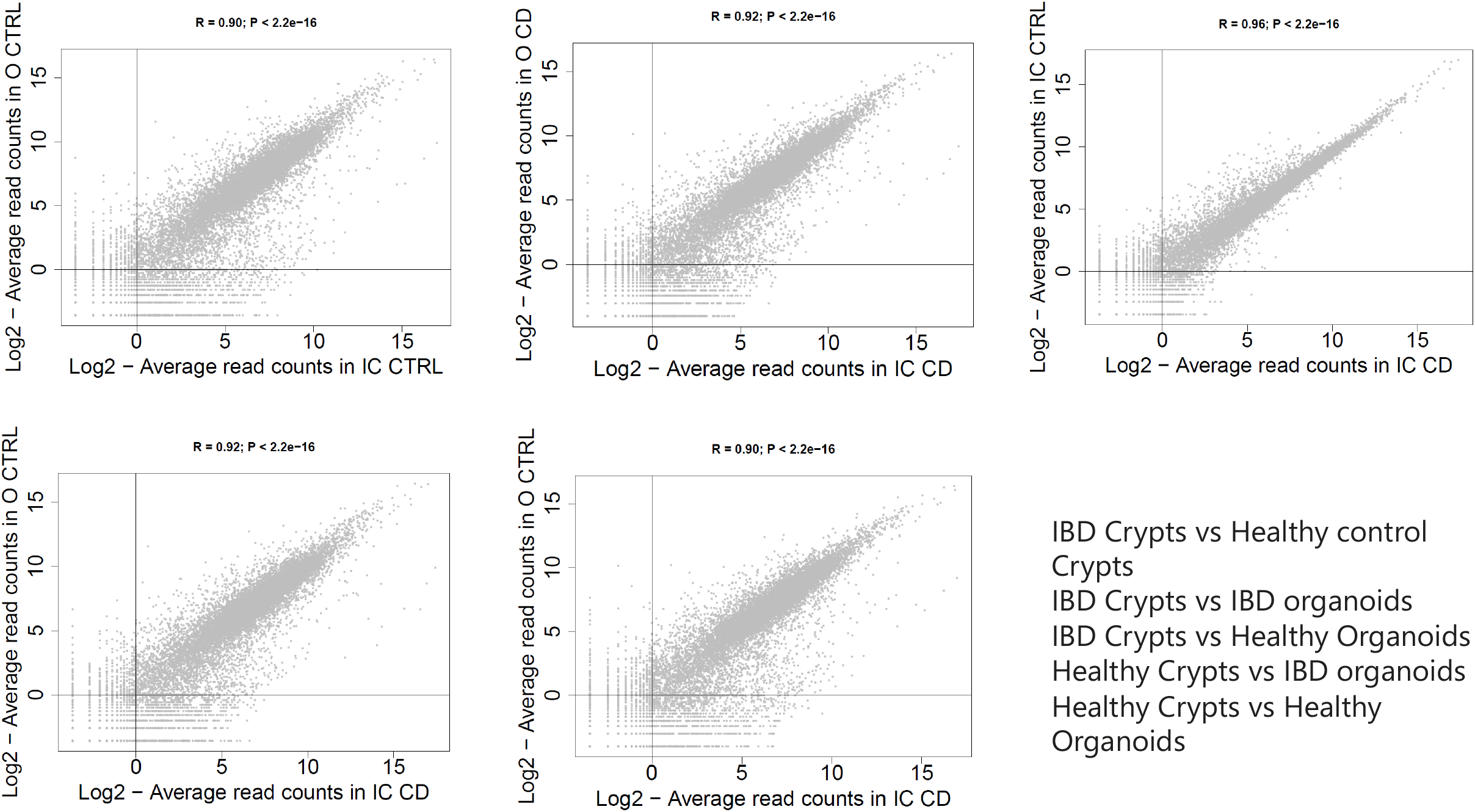

**Supplemental 2.**
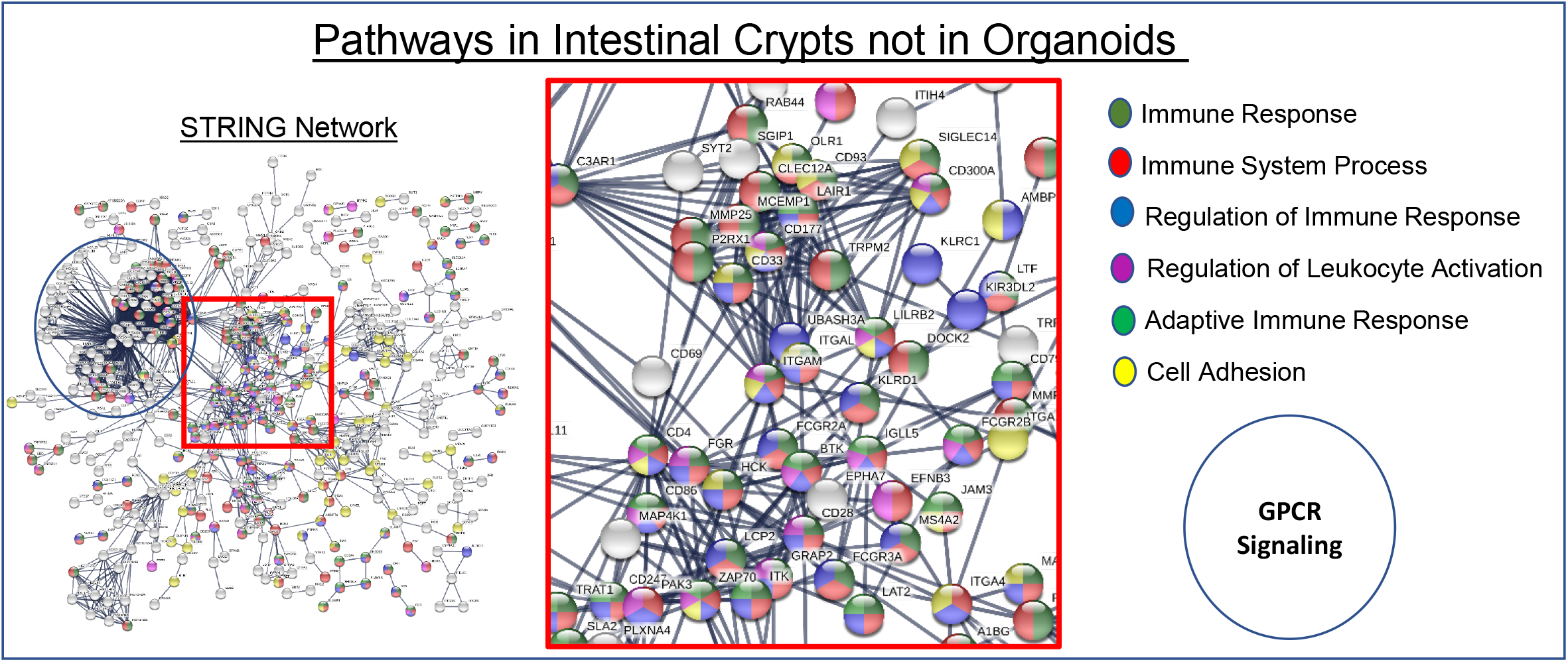

**Supplemental 3.**
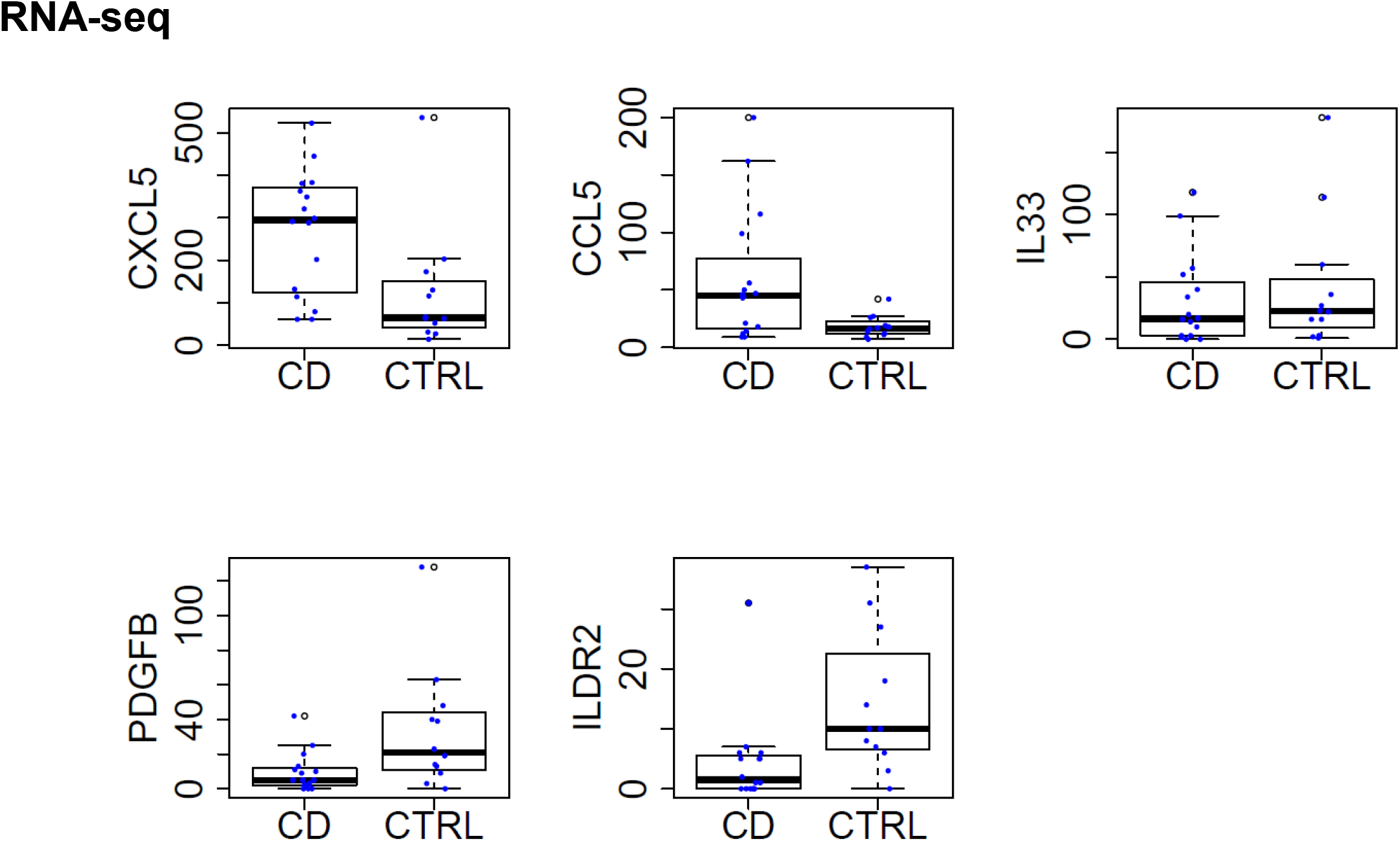

**Supplemental 4.**
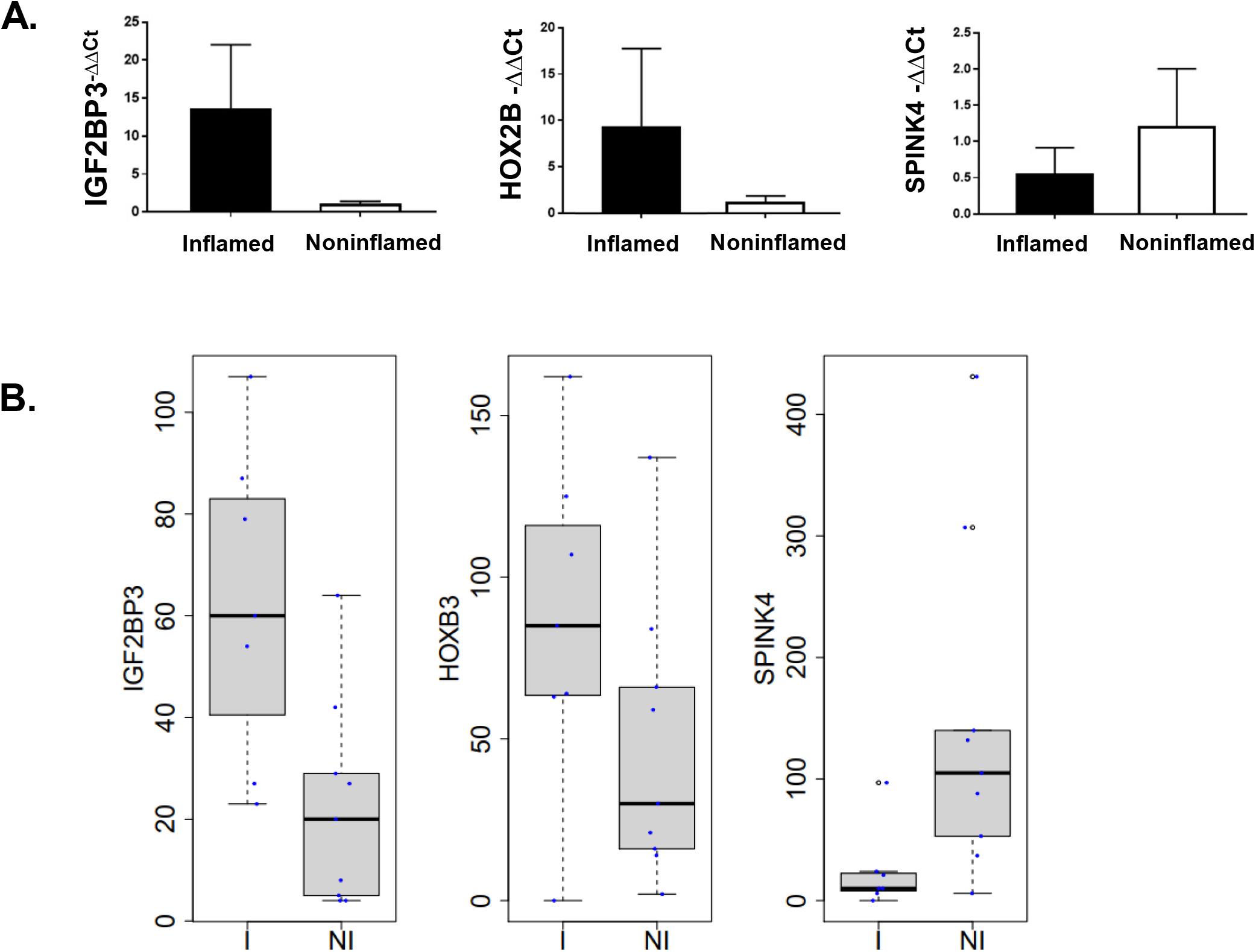

**Supplemental 5.**
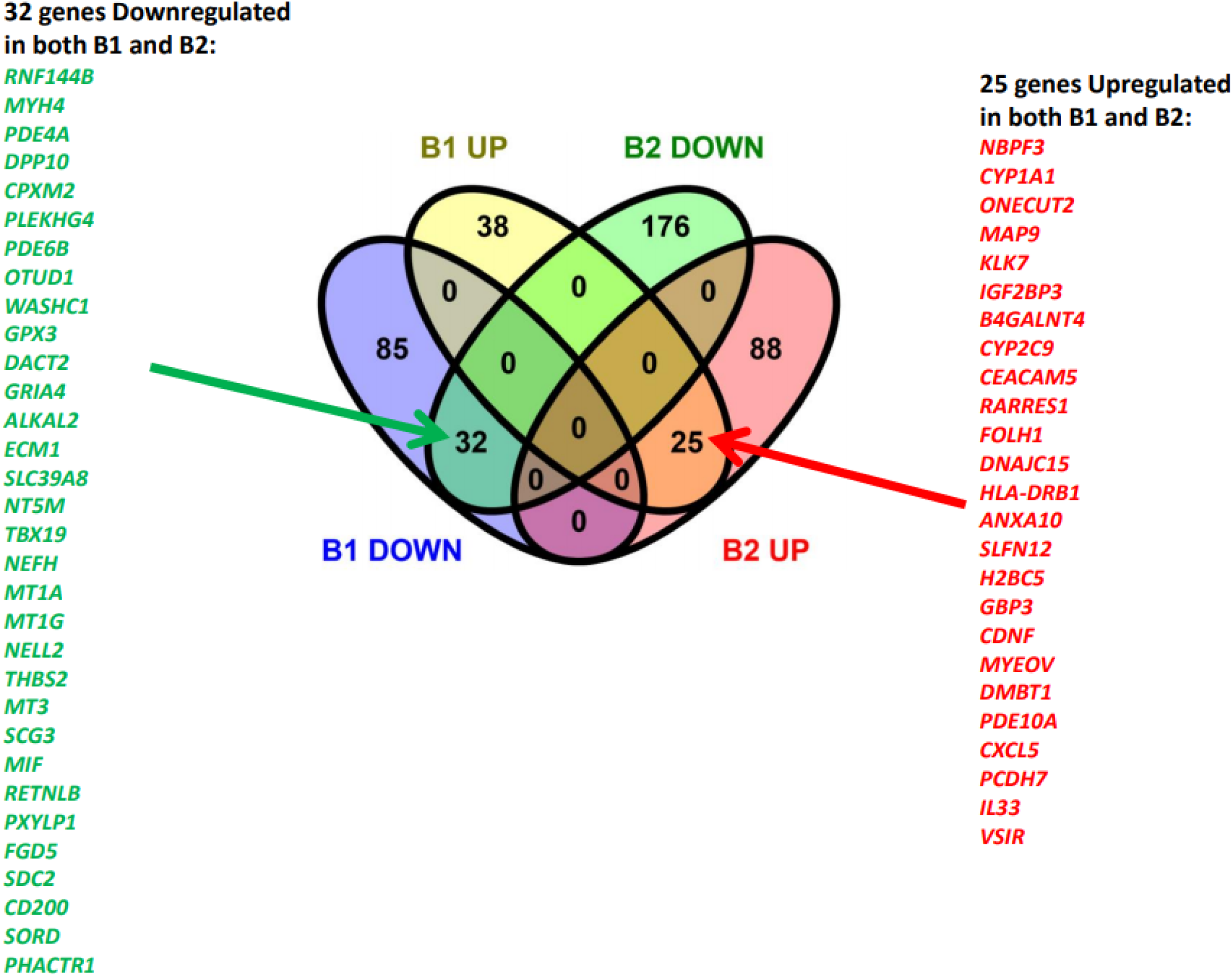

**Supplemental 6.**
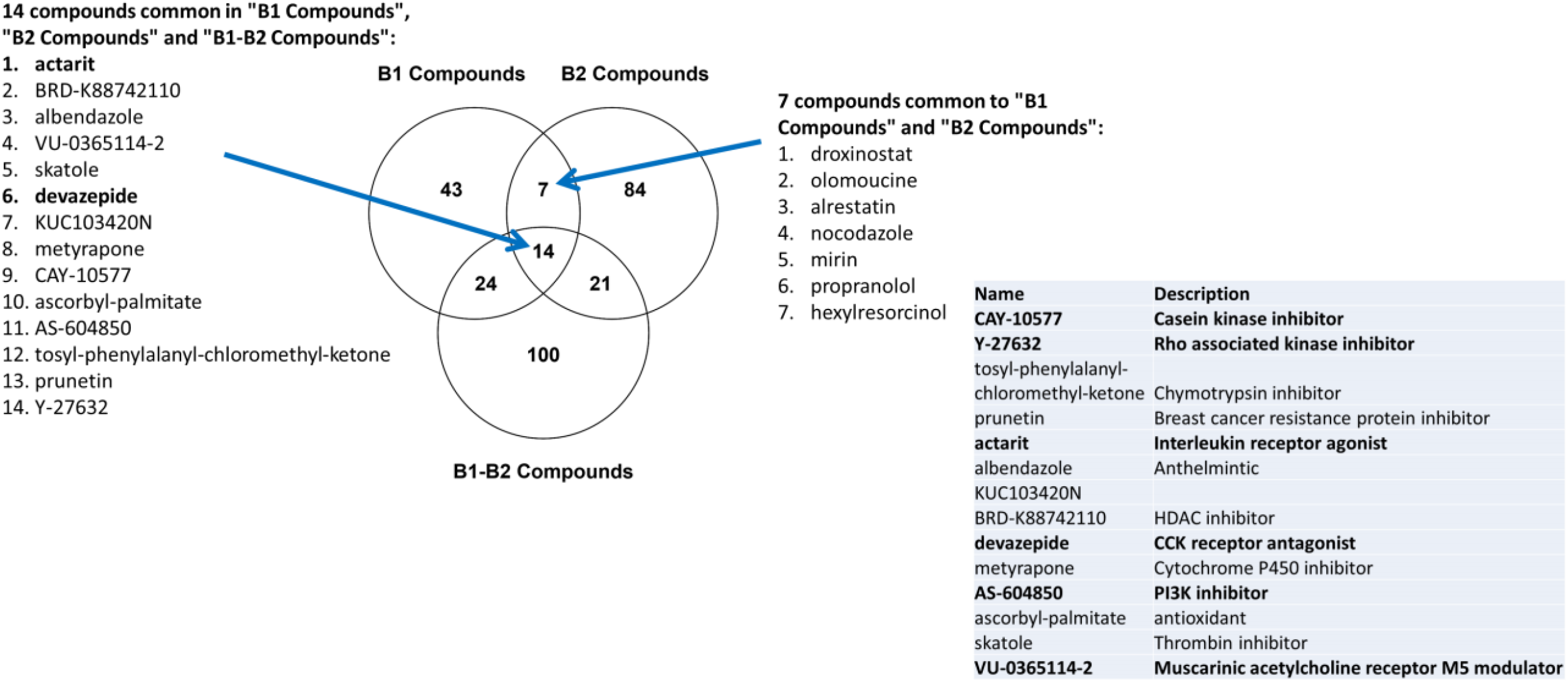

**Supplemental 7.**
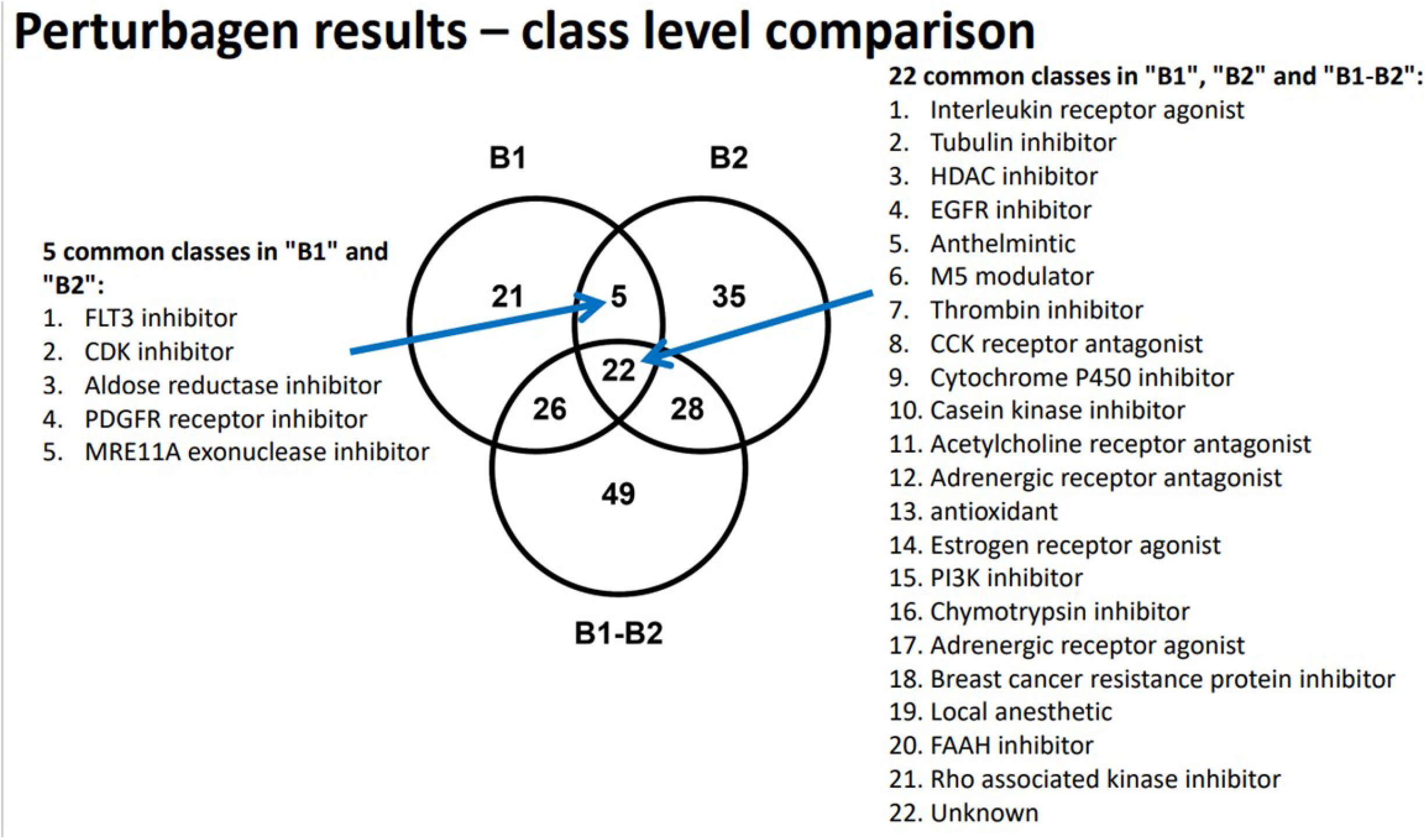

