## Supplementary material for "Ileal derived organoids from Crohn’s disease patients show unique transcriptomic and secretomic signatures": Supp Fig legends

**Transcriptomic signatures from patient driven ileal organoids retain disease states and phenotypes in pediatric Crohn’s disease**

Barbara Joanna Niklinska-Schirtz^1^, Suresh Venkateswaran^1^, Murugadas Anbazhagan^1^, Vasantha L. Kolachala^1^, Jarod Prince^1^, Anne Dodd^1^, Raghavan Chinnadurai^2^, Gregory Gibson^3^, Lee A. Denson^4^, David J. Cutler^5^, Anil G. Jegga^6^, Jason D. Matthews^1^ and Subra Kugathasan^1,2^ *

**Supplementary Information**

**Supplementary Table 1:** The differentially expressed genes (DEGs) identified between CD (n=16) vs non-IBD controls (n=12) in PDOs.

**Supplementary Table 2:** The differentially expressed genes (DEGs) identified between B1 (n=8) vs B2 (n=8) in PDOs.

**Supplementary Table 3:** The differentially expressed genes (DEGs) identified between Inflamed (n=7) vs non-Inflamed (n=9) in PDOs.

**Supplementary Table 4:** The differentially expressed genes (DEGs) identified between B1 (n=8) vs non-IBD controls (n=12) in PDOs.

**Supplementary Table 5:** The differentially expressed genes (DEGs) identified between B2 (n=8) vs non-IBD controls (n=12) in PDOs.

**Supplementary Table 6:** The potential druggable targets identified for the DEGs in B1vs controls (, B2 vs controls as well as the common DEGs among those comparisons are provided.

**Supplementary Table 7:** Patient cohort used for organoid’ conditioned media generation and testing.

**Supplementary Figure 1:** Pairwise comparison analysis between CD and non-IBD controls for both ileal crypts and organoids. The average read counts for ~20,000 protein coding genes are plotted in each plot.

**Supplementary Figure 2:** Signaling pathway analysis based on the genes that are expressed only in ileal crypts when compared to PDOs.

**Supplementary Figure 3:** Boxplots comparison between CD versus non-IBD patients show 5 epithelial specific genes involved in the innate immune response to be significantly different in PDOs. Y-axis shows the actual read counts.

**Supplementary Figure 4:** rt-PCR of IGF2BP3, HOX2B and SPINK4 (**A-C**) to confirm 3 FDR significant genes between inflamed and non-inflamed CD, as found by RNA-seq (**D**).

**Supplementary Figure 5:** Venn diagram shows the DE up- and down- regulated genes which are common among B1 vs non-IBD controls and B2 vs non-IBD controls comparisons.

**Supplementary Figure 6:** Venn diagram shows the number of druggable compounds identified through perturbagen analysis for three DEGs sets. List of 14 druggable compounds identified through perturbagen analysis, which is common for three DEGs sets and their descriptions.

**Supplementary Figure 7:** Venn diagram shows the number of druggable compound classes identified through perturbagen analysis for three DEGs sets.
